# Fundamental Limitations of Contact Tracing for COVID-19

**DOI:** 10.1101/2020.12.15.20248299

**Authors:** P. Tupper, S. Otto, C. Colijn

## Abstract

Contact tracing has played a central role in COVID-19 control in many jurisdictions and is often used in conjunction with other measures such as travel restrictions and social distancing mandates. Contact tracing is made ineffective, however, by delays in testing, calling, and isolating. Even if delays are minimized, contact tracing can only prevent a fraction of onward transmissions from contacts. Without other measures in place, contact tracing alone is insufficient to prevent exponential growth in the number of cases. Even when used effectively with other measures, occasional bursts in call loads can overwhelm contact tracing systems and lead to a loss of control. We propose embracing approaches to COVID-19 control that broadly test individuals without symptoms, in whatever way is economically feasible – either with fast cheap tests that can be deployed widely, with pooled testing, or with screening of judiciously chosen groups of high-risk individuals. Only by ramping up testing of asymptomatic individuals can we avoid the inherent delays that limit the efficacy of contact tracing.

---

The effectiveness of contact tracing for any infectious disease is limited by how quickly contacts can be informed. If contact tracing teams reach an individual’s contacts only toward the end of their infectious period, very few further infections will be prevented. Several delays in the process make rapid contact tracing challenging: the time to develop symptoms, to seek a test, to get test results, and for contact tracing teams to reach contacts. Contact tracing is particularly challenging for COVID-19 because transmission often occurs before symptoms appear [10] and some individuals can transmit who never develop symptoms at all [5].

Symptomatic testing followed by contact tracing, set in the context of ongoing widespread distancing measures, have been the primary means of controlling COVID-19 in many jurisdictions in Europe, the UK and North America. However, broad and restrictive distancing measures have been considered too costly to be palatable in the long term, both economically and in terms of unintended consequences for public health, mental health and inequality [4, 8, 16]. This left most of North America, Europe and the UK, among others, in the difficult position of re-opening their economies following declines in COVID-19 numbers. This reopening has occurred in a context where COVID-19 immunity was very low, substantial costs had been incurred, but COVID-19 had not been eliminated and/or was continually re-introduced. In many areas, contact tracing capacity was dramatically increased to allow reopening.

Since reopening, almost all jurisdictions in North American and Europe have seen substantial resurgences of COVID-19 despite having testing and contact tracing in place. Some have overwhelmed their health care systems, for example exceeding ICU capacity, cancelling elective surgeries, diverting patients, and being depleted of nursing staff, leading in some cases to health care workers being asked to work while testing positive for COVID-19 [11, 17, 18]. Groups of doctors have written open letters calling for wider shutdowns while politicians hesitate, knowing the costs and the unintended damages that these shutdowns will create [1–3]. This has occurred despite symptomatic testing and contact tracing being in place. It is clear that the level of widespread distancing that is tolerable and sustainable is insufficient for robust COVID-19 control, with the testing and contact tracing systems currently in place.

Under normal patterns of social interaction with no contact tracing, *R*_0_ for COVID-19 is estimated to be between 2 and 5 [14]. Social distancing and other NPIs without the use of contact tracing reduce the basic reproductive number *R*_0_ to some lower value *R*^NCT^, the basic reproductive number with no contact tracing. Contact tracing reduces this further to *R* = (1 − *ρ*)*R*^NCT^, where *ρ* is the fraction of cases a contact would infect that are prevented by contact tracing. This fraction depends on the capacity to contact trace and on the timing of symptoms and transmission. Two important features are the fraction of contacts that can be reached and who self-isolate (we call this fraction *α* or “coverage”) and the time from a positive test until the contacts are reached (we call this time *τ*). When cases are rare, contact tracers can rapidly reach a large proportion of a cases’ contacts. As the number of cases rises, contact tracing capacity can become increasingly taxed, reducing the ability to reach all contacts and lengthening the delay. In Figure 1 we illustrate the fraction of cases that contact tracing can prevent given the current tracing capacity, i.e. how *ρ* depends on *α* and *τ*.

**Fig 1.**
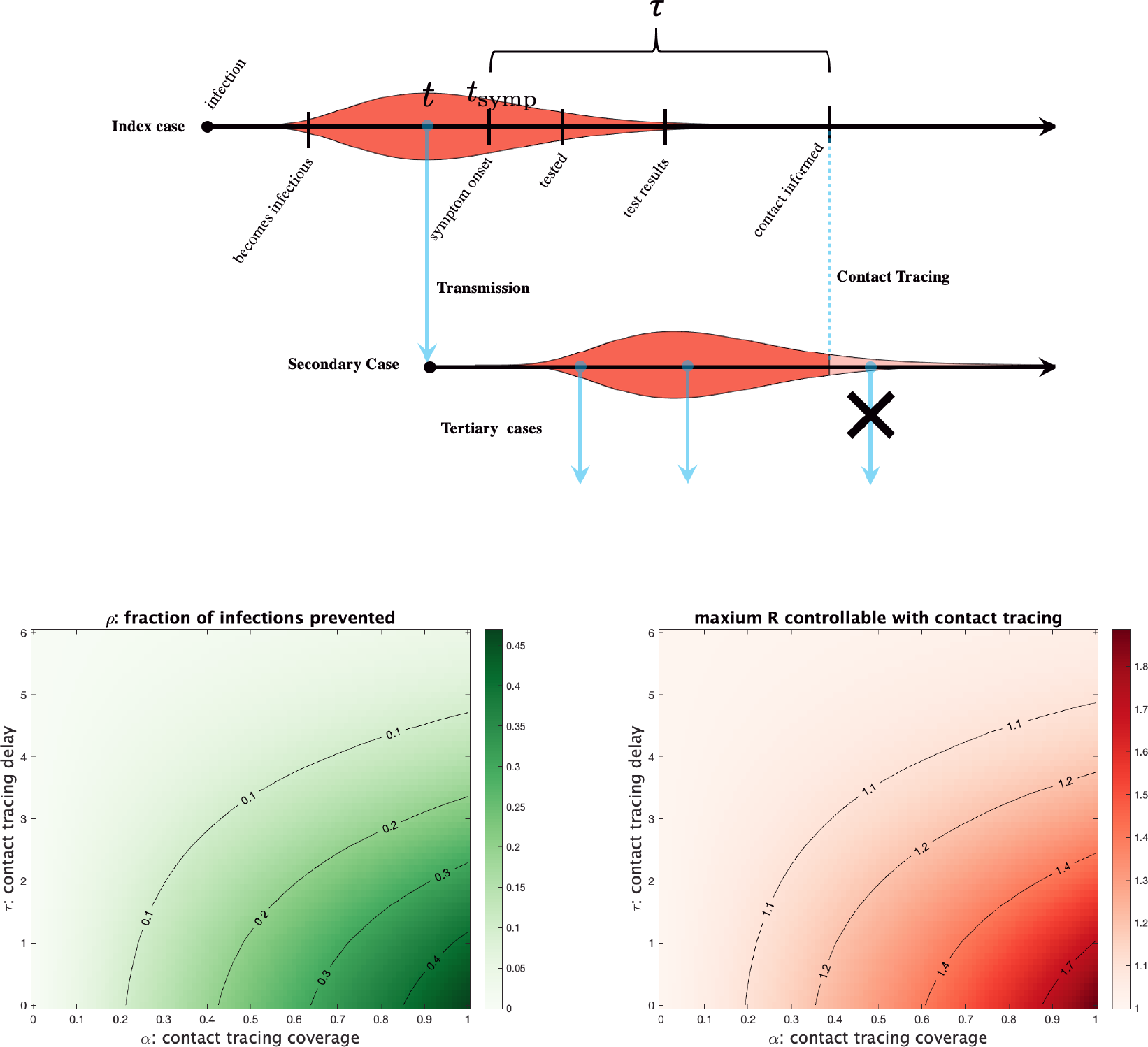
**Top:** Contact tracing of an index case eliminates infections from a secondary case only after contact and self-isolation occur (blue dashed line). **Bottom left:** *ρ* as determined by *α* and *τ*. **Bottom right:** The maximum value of *R*^NCT^ (basic reproductive number without contact tracing) for which contact tracing with a given *α* and *τ* can bring growth under control. **Parameters and calculations:** If a proportion *α* of contacts are reached and self-isolate, the fraction of tertiary infections averted (pink shading) is 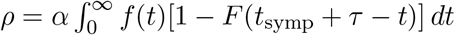 where *f* (*t*) is the distribution of the infectious period (red/pink curve; assumed to be Gamma distributed with a mean of 4 days and standard deviation of 1.5 days [9]) and *F* (*t*) is its cumulative distribution. *ρ* is calculated by assuming that the secondary case started at a random time *t* while the index case was infectious (integrating over *f* (*t*)) and then calculating what fraction of the infectious period of the secondary case is averted (the 1 − *F* term), accounting for the time delay until the index case develops symptoms (*t*_symp_ ≈ 5 days from infection [13]) and their contacts are traced (*τ*).

If we already have contact tracing in place, we cannot observe *R*^NCT^. But if we have estimates of *R*, the coverage and the delay, we can estimate *R*^NCT^ from *R*^NCT^ = *R/*(1 − *ρ*). Then we can determine the fraction of new cases (*ρ*_crit_) we would have to prevent to bring *R* below 1 by solving 1 = (1 − *ρ*_crit_)*R*^*NCT*^. As an example, in British Columbia, on Nov 1, 2020, we estimate *R* = 1.22 ((1.13,1.35) 90% range), with that *τ* approximately 5 days in ideal cases (roughly 1 day for symptom onset to test, 2 days for the test result to be available, and another 2 days for the contacts to be notified, though this varies from one to many days [7]). With coverage of *α* = 0.5, our formula yields *ρ* = 0.08 and *R*^*NCT*^ = 1.33: contact tracing only prevents 8% of onward cases. To reduce *R* to 1 we would need *ρ* = 1 − (1*/*1.33) = 0.25. This could be achieved by reducing delays in contacting from 5 to 2.8 days. Alternatively we could increase coverage, but *R* is only reduced to 1.12 even if we reached 100% of contacts (*α* = 1), which wouldn’t bring spread under control. In reality, a combination of decreasing delays (*τ*) and increasing coverage (*α*) would be optimal.

The fact that there is a critical value of the contact tracing delay beyond which contact tracing is not able to prevent a sufficient fraction of cases to bring COVID-19 under control is an example of a *tipping point*: a value of a parameter where a system has qualitatively different behaviour when the parameter is above or below it [12]. If the delay *τ* is above its critical value, we have exponential growth in the number of cases; otherwise cases decline. There is another tipping point for the coverage; increasing coverage (for example by expanding the definition of a contact to include more people, taking extra measures to insure compliance with self-isolation) could push *R* below 1 if *τ* is sufficiently short and other measures are in place.

Without extensive distancing, contact tracing with realistic parameters will not be able to bring *R* below 1 and we will have exponential growth in the number of cases. But the situation will become worse even for initially very mild exponential growth. As the number of incident cases increases, the contact tracing system will be put under a heavier load, with both *τ* increasing and *α* decreasing, meaning an even lower fraction of cases are prevented by contact tracing. *R* increases even further, which in turn causes even faster growth in the number of incident cases. This positive feedback cycle unabated will lead to the contact tracing system being overwhelmed, since contact tracing capacity cannot be expanded as quickly as case numbers rise.

If a region does have COVID cases under control (*R <* 1), we can also determine how large an increase in call load can be handled before contact tracing will fail to limit spread. As the number of individuals who need to be contacted daily (*n*) rises above the tracing capacity (*c*), the best strategy is to reduce coverage without also adding further delays. In this case, coverage will decline in proportion to how far over capacity the call demand is (e.g., coverage will drop by half if twice as many people need to be called as can be called, *n/c* = 2). Solving for the call volume that will cause *R* to rise above one, we find that contact tracing breaks down once *n/c > ρR*^*NCT*^ */*(*R*^*NCT*^ − 1) or, equivalently, *n/c >* (*R*^*NCT*^ − *R*)*/*(*R*^*NCT*^ − 1). For example, in a region brought down to *R* = 0.9 from *R*^*NCT*^ = 1.33, contact tracing will fail to prevent spread once the demand requires 33% more calls than can be placed in a day.

This is the best case scenario - only coverage was impacted. In reality, as the incidence or call volume rises, delays grow. Testing backs up so that fewer onward cases can be prevented. Thus, any event – a burst in transmission caused by a superspreading event, a cluster of importations, or changes in behaviour around a holiday – that causes the call volume to rise above this limit cannot be reversed by contact tracing alone. Importantly, the number of calls needed can rise either because of true increases in incidence or due to increases in contacts per case – either can cause this kind of collapse. When cases rise, the effectiveness of contact tracing declines just when it is most needed.

Where does this leave us? Heading into mid-winter, with relatively little immunity yet built up and months until the newly approved vaccines become widely available, many jurisdictions must face up to the fact that they cannot have what they so wanted – considerable reopening to avert the high costs of shutdowns – if their primary COVID-19 controls are based on symptomatic testing and contact tracing. Either strong distancing measures must be maintained until a vaccine has been widely deployed, or we must rethink our testing and tracing approach. One option for considerably strengthening the power of contact tracing is to go beyond merely instructing contacts to isolate, but to test all contacts of a known case as rapidly as possible, whether or not they are symptomatic. This approach, which is used in New Zealand [15], has the advantage that if a secondary contact tests positive, tracing for their contacts can be initiated much earlier than if testing only occurs after symptom onset, which might never happen if the secondary contact remains asymptomatic. In contrast, if we simply ask individuals to self-isolate, first this isolation is imperfect, and we do not obtain information about their contacts early enough to prevent onward transmission from them.

Mass testing is another approach recently used in Slovakia, where two thirds of the country were tested with rapid antigen tests over two days. 57,500 COVID-19 cases were identified, which is almost three quarters the number of cases discovered by PCR tests in that country since the beginning of the pandemic [19]. These tests are less accurate than standard PCR tests, but lower cost means that they can be deployed much more widely. Pooled sample testing is another approach: samples are collected from a group and tested at once, reducing the costs of testing [6]. A positive result can lead to instructions to isolate for the whole group and/or to subsequent individual tests. In addition, when call load exceeds contact tracing capacity in a region, mass testing and isolation of positive cases could be used to bring case numbers down to where contact tracing would become effective again. What these have in common is that they aim to find cases, and even contacts of those cases, before symptom onset, at the start of or before infectiousness (effectively reducing *τ*). Measures initiated by testing symptomatic individuals cannot “get ahead of transmission” in the same way.

We can either find ways to get ahead of transmission or continue to use symptomatic testing followed by contact tracing as our primary COVID-19 control. But we now know that contact tracing that focuses on symptomatic cases must be complemented with long, sustained and widespread distancing measures, and these have extremely high economic, social and health costs. If we want to avoid continual shutdowns and resurgences, we need to build robust testing strategies and capacity that can stop transmission much earlier.

## Data Availability

There is no data referred to in the manuscript.

## Notes

### Competing Interest Statement

The authors have declared no competing interest.

### Funding Statement

The authors were supported by Natural Sciences and Engineering Research Council (Canada) grants and by a Genome BC grant. No other external funding was received.

### Author Declarations

N/A - modelling only

## References

1. Covid-19: An open letter to the UK’s chief medical officers. https://blogs.bmj.com/bmj/2020/09/21/covid-19-an-open-letter-to-the-uks-chief-medical-officers/, September 2020. Accessed: 27-11-2020.

2. Shut Down, Start Over, Do it Right. https://uspirg.org/resources/usp/shut-down-start-over-do-it-right, November 2020. Accessed: 27-11-2020.

3. Darren Bernhardt. ‘We are in grave peril’: Manitoba doctors call for emergency funding to deal with COVID-19 spike. CBC News, November 2020. https://www.cbc.ca/news/canada/manitoba/doctors-letter-manitoba-shutdown-covid19-1.5785979. Accessed on 27.11.2020.

4. Giovanni Bonaccorsi, Francesco Pierri, Matteo Cinelli, Andrea Flori, Alessandro Galeazzi, Francesco Porcelli, Ana Lucia Schmidt, Carlo Michele Valensise, Antonio Scala, Walter Quattrociocchi, and Fabio Pammolli. Economic and social consequences of human mobility restrictions under covid-19. Proceedings of the National Academy of Sciences, 117(27):15530–15535, 2020.

5. Oyungerel Byambasuren, Magnolia Cardona, Katy Bell, Justin Clark, Mary-Louise McLaws, and Paul Glasziou. Estimating the extent of true asymptomatic covid-19 and its potential for community transmission: systematic review and meta-analysis. Journal of the Association of Medical Microbiology and Infectious Disease Canada, 2020.

6. Brian Cleary, James A Hay, Brendan Blumenstiel, Maegan Harden, Michelle Cipicchio, Jon Bezney, Brooke Simonton, David Hong, Madikay Senghore, Abdul K Sesay, et al. Using viral load and epidemic dynamics to optimize pooled testing in resource constrained settings. medRxiv, 2020.

7. Penny Daflos. Self-reporting implemented as Fraser Health exceeds contact tracing capacity some days. CTV News, December 2020. https://bc.ctvnews.ca/self-reporting-implemented-as-fraser-health-exceeds-contact-tracing-capacity-some-days-1.5217967 Accessed 12.06.2020.

8. Julie M. Donohue and Elizabeth Miller. COVID-19 and School Closures. JAMA, 324(9):845–847, 09 2020.

9. Tapiwa Ganyani, Cécile Kremer, Dongxuan Chen, Andrea Torneri, Christel Faes, Jacco Wallinga, and Niel Hens. Estimating the generation interval for coronavirus disease (covid-19) based on symptom onset data, march 2020. Eurosurveillance, 25(17):2000257, 2020.

10. Xi He, Eric HY Lau, Peng Wu, Xilong Deng, Jian Wang, Xinxin Hao, Yiu Chung Lau, Jessica Y Wong, Yujuan Guan, Xinghua Tan, et al. Temporal dynamics in viral shedding and transmissibility of COVID-19. Nature Medicine, 26:672–675, 2020.

11. Ivana Kottasová, Gaëlle Fournier, and Niamh Kennedy. The outbreak is so bad in Belgium, some Covid-positive health workers are being asked to keep working. CNN, October 2020. https://www.cnn.com/2020/10/27/europe/belgium-coronavirus-hospitals-intl/index.html. Accessed on 12.06.2020.

12. PJ Lamberson, Scott E Page, et al. Tipping points. Quarterly Journal of Political Science, 7(2):175–208, 2012.

13. Stephen A Lauer, Kyra H Grantz, Qifang Bi, Forrest K Jones, Qulu Zheng, Hannah R Meredith, Andrew S Azman, Nicholas G Reich, and Justin Lessler. The incubation period of coronavirus disease 2019 (covid-19) from publicly reported confirmed cases: estimation and application. Annals of internal medicine, 172(9):577–582, 2020.

14. Ying Liu, Albert A Gayle, Annelies Wilder-Smith, and Joacim Rocklöv. The reproductive number of COVID-19 is higher compared to SARS coronavirus. Journal of Travel Medicine, 27(2), 02 2020. taaa021.

15. Ministry of Health. Contact tracing for COVID-19. https://www.health.govt.nz/our-work/diseases-and-conditions/covid-19-novel-coronavirus/covid-19-health-advice-public/contact-tracing-covid-19. Accessed: 29-11-2020.

16. Betty Pfefferbaum and Carol S. North. Mental health and the covid-19 pandemic. New England Journal of Medicine, 383(6):510–512, 2020. PMID: 32283003.

17. Nicholas Reimann. Here’s Where You Can No Longer Get An ICU Bed Because Of Covid. Forbes, November 2020. https://www.forbes.com/sites/nicholasreimann/2020/11/14/heres-where-you-can-no-longer-get-an-icu-bed-because-of-covid/?sh=58dc15707730 Accessed on 12.06.2020.

18. Laura Romero. As COVID-19 cases rise, some hospitals are again forced to suspend elective surgeries. ABC News, December 2020. https://abcnews.go.com/Health/covid-19-cases-rise-hospitals-forced-suspend-elective/story?id=74492165. Accessed on 12.06.2020.

19. James Shotter. Slovakia’s mass coronavirus testing finds 57,500 new cases. Financial Times.

